# Frequency of Hypoglycemic Episodes in Patients with Type 2 Diabetes After Medication Changes at Hospital Discharge: A Retrospective Cohort Analysis

**DOI:** 10.1101/2025.08.11.25333413

**Authors:** Avraham Ishay, Nimra Maman, Elena Chertok Shacham

## Abstract

Hypoglycemia is associated with an increased risk of emergency room visits, rate of hospitalizations, and lower quality of life. Recommendations given to patients with diabetes mellitus at hospital discharge provide an opportunity to improve long-term glycemic control but may increase the short-term risk of hypoglycemia.

In this retrospective study, we investigated the hypoglycemic events occurring within three months following hospitalization for common medical conditions in 693 patients with type 2 diabetes. We examined whether the recommendations for diabetes treatment given at discharge are linked with these events.

We found that patients recommended to modify their home diabetes treatment had a higher rate of hypoglycemia of any severity than those discharged without change (p=0.003). We conclude that individualized discharge treatment plans, frequent follow-ups, and continuous glucose monitoring for high-risk patients are recommended to prevent hypoglycemic events.

## Introduction

Hypoglycemia is a common complication in both type 1 and type 2 diabetes (T2D) and a limiting factor in treatment (1).

Predictors of hypoglycemia in T2D patients include insulin treatment duration, previous hypoglycemic episodes, comorbidities, aging, renal function decline, and cognitive impairment. (2). Patients experiencing hypoglycemia or maintaining near-normal glucose levels while hospitalized face an elevated likelihood of hospital readmission and mortality both during their hospital stay and after discharge (3,4). Intensifying diabetes treatment at discharge may improve outpatient diabetes control post-hospitalization but may be associated with an increased short-term risk of severe hypoglycemia in older adults (5, 6). The transition period following hospital discharge is a high-risk period for patients with diabetes with an increased risk of both severe hyperglycemia or hypoglycemia and readmission (7). Tailored discharge plans, including medication reconciliation, are recommended to prevent post-discharge complications. (8).

## Methods

This retrospective cohort study, conducted at HaEmek Medical Center in Afula, Israel, examined electronic health records (EHR) of T2D patients admitted to internal medicine departments from 2013 to 2020. Inclusion criteria were patients (aged 18-90) treated with insulin during hospitalization who experienced hypoglycemia within three months post-discharge.

Hypoglycemia was categorized by severity: severe (glucose <50mg/dL), moderate (two or more events with glucose 50-70mg/dL) and mild (one event with glucose 50 70 mg/dL), based solely on laboratory results.

We compared hypoglycemic events in patients whose diabetes treatment was modified after discharge and those whose treatment remained unchanged.

Change in diabetes treatment was defined as a change in insulin dosage, addition of new insulin, or a new class of drugs.

Regimens were classified into four groups:

1. Insulin only
2. Insulin with oral medications
3. Insulin plus Glucagon-like peptide 1 receptor agonists (GLP1ra)
4. Combination therapy

Mortality was assessed at 90- and 365 days post-hospitalization.

An increase of more than 50% of the basal serum creatinine was considered as an acute renal failure.

Multivariate ordinal regression analyzed the severity of hypoglycemia, while linear regression estimated the initial hypoglycemic values. To mitigate the influence of potentially confounding variables, including age, comorbidities, and duration of hospitalization, we implemented multivariate statistical analysis. Linear regression was chosen to estimate the value of the initial hypoglycemic event. It was particularly suitable as all participants in the study experienced at least one hypoglycemic event, making logistic regression less appropriate. Ordinal regression was applied to assess the severity of hypoglycemia, considering two key factors: the number of hypoglycemic events and the glucose values associated with these events. Both models were adjusted to reflect the continuous value of the first hypoglycemia measurement and the ordered severity of subsequent hypoglycemic events.

### Statistical analysis

Standard distribution measures (mean, standard deviation, and median) were presented for both groups (change/no change in treatment).

Frequencies (%) were presented for categorical variables by groups. Additionally, the association between different variables was calculated using a chi-square or non-parametric Fisher’s exact test. Comparison between groups was made using independent samples t-test and paired t-test. Multivariable analysis was conducted using ordinal regression to predict the severity of hypoglycemia (adjusted odds ratio) and linear regression to predict the value of the first hypoglycemic event after discharge.

Data analysis was performed using IBM SPSS Statistics V 24 software. Statistical significance will be considered when *P*-value<0.05.

## Results

Of 693 patients analyzed, 510 had changes made to their diabetes treatment upon discharge, while 183 continued their preadmission treatment. Patients with medication modifications at discharge exhibited a higher incidence of hypoglycemic events post-discharge compared to those without change. Eighty-four percent of severe hypoglycemic events occurred in patients who had a change in treatment at discharge (p<0.003, Table 1). Multiple severe events were also more frequent in patients who had medication changes compared with those who did not (78.9% vs. 21.1%). For moderate hypoglycemic events, the figures were similar, with 76.3% of events occurring in patients with changes versus 23.7% for those without changes. Mean blood glucose levels were similar in both groups: 183.9±73.5mg/dL (no change) vs. 185.4±74.3mg/dL (with change), p=0.86. Overall, there was an improvement in diabetes control, with HbA1c levels decreasing from 8.4±1.8% before hospitalization to 8±1.8% within 3-6 months after discharge (P<0.0001). The reduction in HbA1c was consistent across both study groups.

**Table 1.** Clinical and demographic characteristics of two groups of patients.

| **Clinical data** | **Overall**<br>**N= 693** | **Change of**<br>**treatment<sup>†</sup>**<br>**N=510** | **No change of**<br>**treatment**<br>**N=183** | **P-value** |
| --- | --- | --- | --- | --- |
| Age, Mean±SD | 70.8±11 | 70.3±11 | 72±11 | NS |
| Age, Median (IQR) | 71.4 [15] | 71.6 (15.6) | 71.2 (14.6) | NS |
| Male, n (%) | 362 [52%] | 124 (34.3%) | 238 (65.7%) | NS |
| Hypoglycemia <50, n (%) | 136 [19.6] | 114 (84%) | 22 (16%) | 0.003** |
| Hypoglycemia <70, n (%) | 557 [80.4%] | 396 (71%) | 161(29%) |  |
| Insulin treatment, n (%) | 515 [74%] | 373 (72%) | 142 (28%) | NS |
| Insulin dose u/kg Mean±SD | 0.7±1.2 | 0.8±1.4 | 0.6±0.3 | NS |
| Oral treatment, n (%) | 195 [28%] | 158 (81%) | 37 (19%) | 0.005* |
| GLP-1 combination, n (%) | 22 [3%] | 17 (77%) | 5 (23%) | NS |
| HbA1C mg% before, Mean±SD | 8.4±1.8 | 8.5±1.8 | 8.4±1.8 | NS |
| HbA1C mg% after, Mean±SD | 8±1.8 | 8.1±1.8 | 7.9±1.6 | NS |
| GFR (ml/min), Mean±SD | 71.6±48 | 72.4±46.5 | 70±51.3 | NS |
| LOS<sup>‡</sup> >5 days, n (%) | 358 [52%] | 132 (34%) | 226 (66%) | NS |
| LOS 3-5 days, n (%) | 238 [34%] | 74 (31%) | 164 (69%) |  |
| LOS 1-2 days, n (%) | 97 [14%] | 26 (27%) | 71 (73%) |  |

<sup>†</sup> Change of treatment: Insulin dose change: n =281, Drug class change: n =229
<sup>‡</sup> LOS: Length of Hospital Stay
Note: Column percentages are shown in brackets; row percentages are shown in parentheses

Within 90- and 365 days post-discharge, 124 (17.8%) and 319 (46%) patients died, respectively. No significant correlation was found between mortality and changes in treatment plans at discharge nor between mortality and hypoglycemic events. The risk factors for severe hypoglycemia included in the multivariable analysis models were age, gender, comorbidities (cardiovascular disease, chronic kidney disease, congestive heart failure, cerebrovascular disease, dementia), preadmission and discharge insulin doses, hospital length of stay, insulin dose change at discharge, and renal function deterioration during hospitalization (defined as an increase of >50% in serum creatinine).

Patients with chronic renal failure (CRF) had a 2.76 times higher risk of experiencing severe or moderate hypoglycemia compared to those with normal renal function. For each year increase in age, the odds of experiencing severe or moderate hypoglycemic events increased by 16.6 times. Acute renal failure was also associated with an increased risk of post-discharge severe hypoglycemia (Fig. 1). The ordinal regression model, which included the eGFR variable, showed significant improvement in predicting the severity of hypoglycemia outcomes (P<0.001). However, the linear regression model, which added the preadmission oral antidiabetic medications variable, was not significant (P=0.87) in predicting glucose values at the first hypoglycemic event.

**Figure 1.**
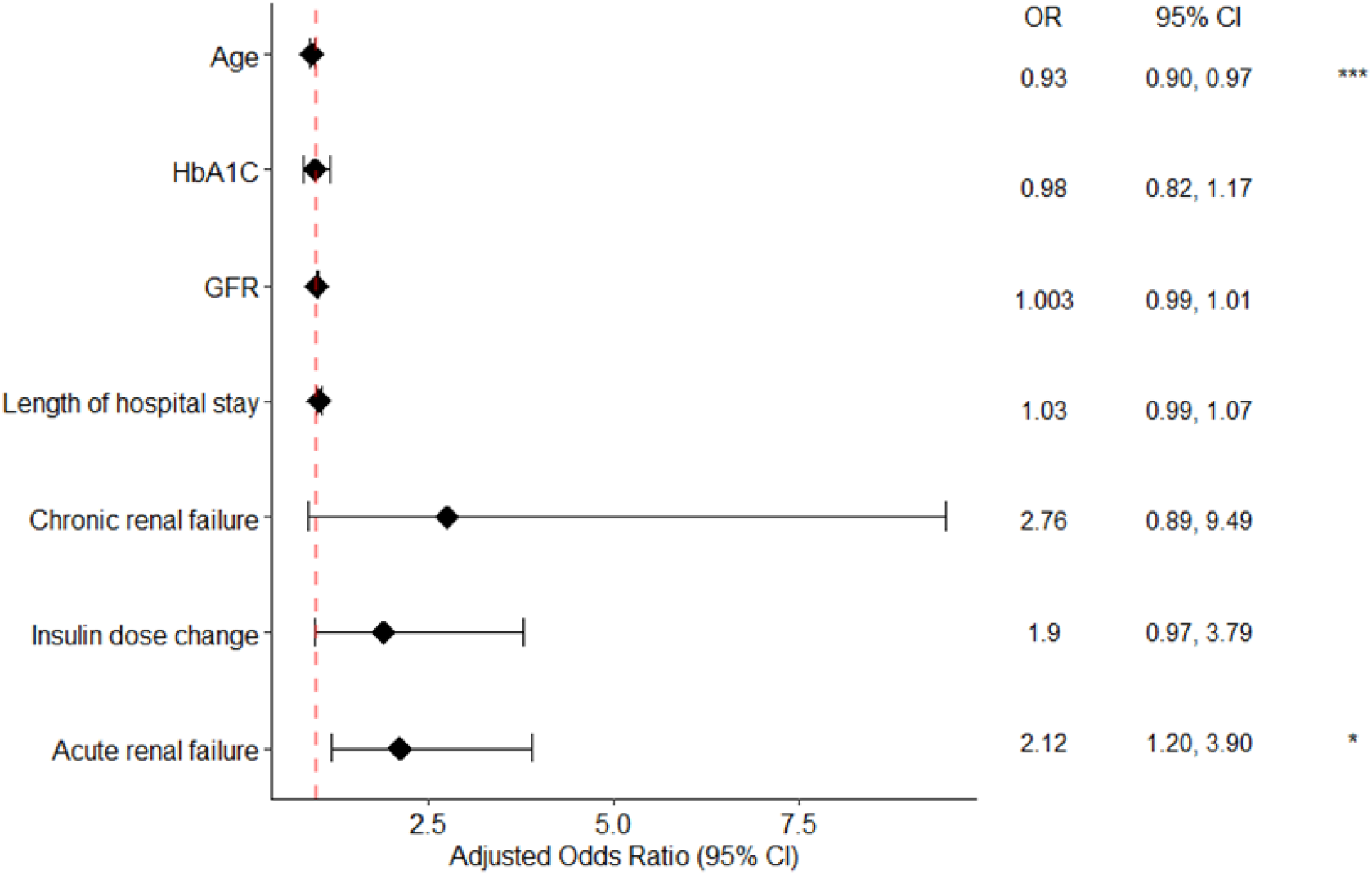
Odds ratio and confidence intervals (CI) for factors associated with severe hypoglycemia presented in a forest plot. Logistic regression model of risk factors associated with severe hypoglycemia presented in a forest plot with odds ratio (OR) and confidence intervals (CI). * *P*<0.05 \*\*\**P*<0.01

## Discussion

Our study found that modifying antidiabetic therapy at hospital discharge increases the frequency and severity of hypoglycemic events in older adults with diabetes within three months, consistent with existing research (9,10). The risk is notably high in patients receiving insulin before hospitalization and at discharge. In our cohort, a high percentage of patients (74%) received insulin before hospitalization. At discharge, insulin use was even higher in both groups (92% and 78% in the change and no change groups, respectively). The risks and adverse consequences of hypoglycemia may be underreported in Randomized Controlled Trials (RCTs) compared with real-world data and are not always clearly appreciated in people with type 2 diabetes (2). In middle-aged and older adults with type 2 diabetes, severe hypoglycemia has been associated with an increased risk of cardiovascular events and mortality (11). In our study, although changing the diabetes medication regimen at discharge led to more hypoglycemic events, it did not increase the one-year mortality rate.

In line with the study by Hung et al. (12), we observed a significant correlation between renal function decline during hospitalization and the severity of post-discharge hypoglycemic events. Potentially unnecessary intensive treatment at discharge, including an increment in insulin dosage, is associated with an increased risk of severe hypoglycemia (13). While we found that post-discharge insulin doses were similar between groups, those with medication changes had higher preadmission insulin doses and a higher risk of severe hypoglycemia, even with lowered post-discharge doses. Novel antidiabetic drugs may offer a better benefit-harm profile post-discharge (14). However, in our study, only a minimal percentage of patients (4%) were discharged with GLP1ra.

Our findings indicate an increased risk of hypoglycemic events following changes in diabetes medication regimens at discharge. However, this should be interpreted cautiously due to the study’s focus on older adults and its limitation to a single-center setting. Furthermore, the reliance on laboratory data to identify hypoglycemic events may lead to the omission of undocumented cases. However, given the number of older adults with diabetes who are hospitalized, the risk of post-discharge hypoglycemic events is significant and has to be taken into consideration.

## Conclusion

Changes in diabetes treatment plans at discharge can increase the risk of severe hypoglycemia in older adults with diabetes. Key risk factors include deterioration of renal function, insulin dosage changes, and preexisting chronic renal failure.

Individualized treatment plans, frequent check-ups, and continuous glucose monitoring for high-risk patients are crucial for prevention.

## Data Availability

All data produced in the present study are available upon reasonable request to the authors

## Disclosure of ethical statements

1. The Institutional Review Board approved the research protocol. Approval number 0050-21 (May 21, 2021)
2. Informed consent: N/A
3. Approval date of Registry and Registration No of the study/trial: N/A: There is no such requirement for retrospective data studies in Israel.
4. Animal studies: N/A
5. Conflict of interest: The authors or their institutions have no financial or nonfinancial relationship to disclose for the submitted manuscript.

